# Ethical and human subject burdens of trials conducted to evaluate biosimilars

**DOI:** 10.1101/2021.03.05.21252938

**Authors:** Jennifer A. Ohn, Preston J. Atteberry, Mark R. Trusheim, Peter B. Bach

## Abstract

**Background/Aims:** With a policy goal of introducing price competition into the market for biologic drugs after their period of market monopoly is over (called ‘loss of exclusivity’), policymakers created a pathway for companies to make copies of those treatments and termed them ‘biosimilars’. But unlike generic drugs, biosimilar drug copies must be studied in human trials to assure they have the same clinical effect as the original biologic products. The burden that this places on human subject participants, and the opportunity cost on the clinical trial system generally, have not been considered in detail.

**Methods:** For all biosimilar drugs in development, approved, or that failed to obtain approval in the US, we abstracted from clinicaltrials.gov registry the number of subjects enrolled at each phase of development.

**Results:** We identified 105 clinical trials for approved or withdrawn biosimilars and another 20 studies that are either planned, ongoing or completed for biosimilars in development. These studies collectively enrolled (or plan to enroll) 38,169 human subjects. Most (28,130) are enrolled in phase 3 studies. The mean number of human subject participants per approval is 1,045, about 25% of the number required for a new drug approval on average.

**Conclusions:** A consequential number of human subjects are required for the testing of biosimilar drugs prior to approval. The explicit and sole purpose of biosimilars is to induce competition in order to lower prices of biologic drugs after loss of exclusivity. The burden the biosimilar approval trials place on human subjects with no direct clinical benefits but definite risks, and the possibility that they rob subjects from trials that are of more scientific importance, are ones policymakers might consider. Price regulation of biologic drugs after loss of exclusivity could achieve lower prices as well, without the burdens of the current approach.

**Funding Source:** Arnold Ventures (Grant to support Drug Pricing Lab at Memorial Sloan Kettering Cancer Center)

---

A central tenet of pharmaceutical pricing policy is that innovator companies only charge monopoly prices for a fixed period. Subsequently prices of their products are expected to fall considerably, and today our system relies on competitive pricing pressure from alternative versions of the original innovator drug to achieve this objective.^1^ People may not be familiar with this policy model per se, but the notion that new drugs get a certain period on the market when their prices are high, and that later other companies make copies of them termed *generic drugs* and prices fall often by a huge amount, is the operational manifestation of that policy. Generic drug entry has accomplished its intended goal in large measure, as today 8 in 10 small-molecule drugs are generics, and prices of innovator drugs fall 85-95% upon entry of multiple generic competitors.^2,3^ The system has been in place since the passage of The Drug Price Competition and Patent Term Restoration Act of 1984 (the Hatch-Waxman Act). One reason, perhaps the central reason, generic drug competition has been so effective, is because small molecule drugs that face generic competition are relatively easy to copy directly as they have straightforward chemical structures. The US FDA allows them on the market without requiring extensive human trials that would test if the generic copies have the same clinical effect, a huge savings to competitors in money, time and risk.

As of about twenty years ago, a newer category of drugs called biologics entered the prescription drug space in force. Huge blockbusters such as Rituxan and Humira belong in the category. Though they even today represent a small subset of prescriptions (around 2%), they account for a large share of spending on pharmaceuticals overall of about 40%.^4^The Biologic Price Competition and Innovation Act (BPCIA) of 2009 created a path of entry for the equivalent of generic drugs called ‘biosimilars’ to enter and compete on price with biologic drugs. But since its passage biosimilar entry has not been nearly as effective. One reason is likely that biologic drugs are impossible to just copy. They are highly complex large molecules that require extensive, expensive, and time-consuming development to reach a point where they constitute a plausible replica of the original biologic drug. The development of a single biosimilar drug typically spans seven to eight years and costs between $100 to $200 million.^5,6^ This high expense reflects the reality that the US FDA only grants approval for a biosimilar when the maker documents the biosimilar works similarly to the original, reference, biologic drug it aims to replicate in extensive human trials. These trials include late stage trials involving individuals affected with the conditions the biosimilar targets.

The requirement for extensive late stage testing of biosimilars prior to approval incurs a toll that has not been well explored or characterized. The toll includes an unmeasured opportunity cost, in that enrolling subjects with conditions on trials of biosimilars likely robs trials of innovative treatments for the same condition of potential human volunteers. But also these trials, like all interventional trials, impose a meaningful cost on human subject participants – both inconvenience and risk. Under the traditional ethical rationale for those burdens placed on subjects in clinical trials, they must be counter-balanced through benefits that the trials may produce. In our view, the counter-balancing of the burdens imposed on enrolled human subjects with possible benefits is the most important principle underlying the ethical conduct of human subjects research. Ethical trials offer volunteers an explicit tradeoff – internalize the risk of testing the experimental drug, your participation will potentially bestow benefits on others through the knowledge the trial generates. (This core tradeoff underlies essentially all acts of volunteerism). The problem is that biosimilar trials have no potential to offer a counter-balancing benefit to others.

We sought to approximate these two dimensions of costs incurred by running trials of biosimilars by counting the number of human research participants biosimilar clinical trials have involved, for both already approved biosimilars and for ones in late stages of development. As of December 20, 2020 (according to clinicaltrials.gov), there have been 102 completed clinical trials that served as the basis of FDA approval of 29 biosimilars, and there are another 20 studies that are completed, ongoing, or planned for as yet unapproved biosimilars (Table 1). The median number of trials registered for all biosimilars is 2. The mean number of subjects per approved biosimilar is 1,045, and the mean number of subjects for biosimilars in an advanced stage of development is 682. All told, completed and ongoing studies involved or anticipate involving 38,169 subjects (Table 1).

**Table 1:** Number of trials and number of involved human subjects in approval studies of biosimilars by Phase.

| Phases of trials | Number of trials | Number of subjects |
| --- | --- | --- |
| Phase I* | 54 | 9,155 |
| Phase II | 4 | 396 |
| Phase III | 65 | 28,130 |
| Phase IV | 2 | 488 |
| Totals | 125** | 38,169 |

\*Category includes both trials listed as Phase 1 and those listed as Phase 1/2
\*\*Represents 102 studies of approved biosimilars, 20 studies of biosimilars in development, and 3 studies for a biosimilar that failed to reach approval
Note: In general, biosimilar approval involves larger early phase trials to evaluate similarity to reference product on pharmacokinetic outcomes than do approvals of new biologic drugs without a history to draw upon<sup>12</sup>

To put that number in some perspective, there were 154,000 participants in trials supporting approval of 34 novel drugs by the FDA in 2015, or around 4,500 subjects per approval.^7^While certainly not all subjects enrolling in trials of biosimilars would be eligible for interventional trials of novel therapies, many might be as both types of studies enroll afflicted patients willing to consent to studies of experimental therapies. As such these totals suggest that the number of subjects potentially diverted from important clinical trials to test biosimilars is consequential, and that ongoing biosimilar trials may be hampering the pace of testing of innovative therapies. Harder to measure, but no less worthy of consideration as a result, biosimilar clinical trials require researcher and investigator time, both themselves limited resources and thus additional dimensions of the opportunity cost imposed by reliance on biosimilar entry and subsequent competition for cost savings. The average time from clinical trial site selection to study start-up is 31.4 weeks, in addition to the six to seven years it takes for the development for a single biosimilar drug.^8^

As for the costs to the human subject volunteers, not only does participation take time, and always require more clinical encounters, tests, and a variety of other additional interactions with health care providers beyond those needed for routine care, but also studies expose subjects to risks. Communicating these risks is in fact the core purpose of the informed consent process, and risks are always considered significant when trials involve interventions such as drug therapy. In the case of biosimilar human testing, those risks come largely from the possibility that the biosimilar will not be as effective, or as safe, as the innovator molecule it seeks to displace. For instance, novel drug-related side effects were observed with the introduction of an early biosimilar erythropoietin product in Thailand. ^9,10^ This risk is one of several reasons why human trials of biosimilars are required prior to marketing approval. There can also be manufacturing process issues that affect new products from time to time. In 2013 Takeda recalled Omontys (peginesatide), an erythropoietin agent, in response to reports of hypersensitivity due to minute manufacturing variability.^11^ Johnson and Johnson instituted a similar recall for Eprex (epoetin alfa).^12^

In traditional human subjects research, reasonable risks to the participants are acceptable because they are counter-balanced by the benefits of the knowledge the research produces. Whether it uncovers benefits or no new benefits of a new treatment, it still adds to our collective knowledge. But in the case of clinical trials of biosimilars, the very nature of the endeavor requires that the risks are not offset by knowledge gains that can benefit others. Rather, biosimilar development is titrated to achieve equivalence. Evidence of clinical superiority of a biosimilar product would prevent FDA approval as a biosimilar in fact. The BPCIA states that approval of a biosimilar should be based on evidence of “no clinically meaningful differences” relative to the reference product.

Because biosimilar trials are not designed to potentially improve clinical benefits for patients, they fall short of the ethical standards of clinical research. The United States Clinical Trial Registry website states that clinical studies are “intended to add to medical knowledge,” stipulating that risks of participation in clinical trials “may not be greater than the risks related to routine medical care.” The World Medical Association’s Declaration of Helsinki states that “the primary purpose of medical research is to generate new knowledge,” also noting that this goal “can never take precedence over the rights and interests of individual research subjects.”^13^

We can anticipate some counter arguments to our conclusion that trials of biosimilar treatments impose costs on the pace of innovation and unacceptable costs (due to lack of counter-balancing benefits) on human subject volunteers. Some may argue that future patients will indeed benefit from cost savings that approved biosimilars will generate. Perhaps the ethical justification of clinical trials could be broadened with respect to what benefits justify exposing subjects to risk, and thereby incorporate cost savings as a benefit. But to our knowledge at present the paradigm is generally restricted to harms and benefits related to health itself. This objection prompts a second question to which we think we can offer an answer. Namely, are we trapped in a model where we must introduce biosimilar competition in order to see lower prices of biologics once they have lost their exclusivity? We do not think so. Policymakers could instead price regulate biologic drugs at that point, just as they price regulate hospital and physician fees in Medicare and price regulate electric utilities. We recently proposed such a solution to address the reality that biosimilar competition has not been effective at lowering prices.^14^

A second counter to our concern might be that the subjects enrolling in these trials are not really being exposed to additional risks, based on a supposition that the biosimilar products being evaluated are basically the same as the innovator molecule. But if this were known with certainty prior to the evaluation of these products, then evaluations would not be required. A biosimilar is not Schrodinger’s Cat, it cannot simultaneously be known to be as effective and safe as the innovator molecule it is designed to replicate and require testing to see if it is so.

The unnecessary and substantial opportunity and human subject costs imposed by required human testing of biosimilar drugs is a source of ethical concern and economic inefficiency, and these should be counted against the payer benefits of biosimilars.

## Data Availability

The datasets generated during the current study are available from the corresponding author on reasonable request.

## Appendix

**Number of patients enrolled in biosimilar clinical trials**

| Brand Name of Biosimilar Drug | Generic Name | No. of Participants Enrolled in Completed Trials | No. of Participants Enrolled in Ongoing Trials | No. of Participants Enrolled in All Trials |  |
| --- | --- | --- | --- | --- | --- |
|  |  |  |  | N | % |
| Approved Biosimilar Drugs |  |  |  |  |  |
| Mvasi | bevacizumab-awwb | 892 | 170 | 1,062 | 2.8% |
| Zirabev | bevacizumab-bvzr | 821 |  | 821 | 2.2% |
| Erelzi | etanercept-szsz | 907 |  | 907 | 2.4% |
| Eticovo | etanercept-ykro | 834 |  | 834 | 2.2% |
| Retacrit | epoetin-epbx | 1,615 |  | 1,615 | 4.2% |
| Herzuma | trastuzumab-pkrb | 1,015 | 174 | 1,189 | 3.1% |
| Kanjinti | trastuzumab-anns | 882 |  | 882 | 2.3% |
| Ogivri | trastuzumab-dkst | 632 |  | 632 | 1.7% |
| Ontruzant | trastuzumab-dttb | 984 |  | 984 | 2.6% |
| Trazimera | trastuzumab-qyyp | 1,200 |  | 1,200 | 3.1% |
| Abrilada | adalimumab-afzb | 1,333 | 420 | 1,753 | 4.6% |
| Amjevita | adalimumab-atto | 1,343 |  | 1,343 | 3.5% |
| Cyltezo | adalimumab-adbm | 2,629 |  | 2,629 | 6.9% |
| Hadlima | adalimumab-bwwd | 972 |  | 972 | 2.5% |
| Hulio | adalimumab-fkjp | 1,937 |  | 1,937 | 5.1% |
| Hyrimoz | adalimumab-adaz | 818 |  | 818 | 2.1% |
| Fulphila | pegfilgrastim-jmdb | 411 |  | 411 | 1.1% |
| Nyvepria | pegfilgrastim-apgf | 575 |  | 575 | 1.5% |
| Udenyca | pegfilgrastim-cbqv | 541 |  | 541 | 1.4% |
| Ziextenzo | pegfilgrastim-bmez | 624 |  | 624 | 1.6% |
| Nivestym | filgrastim-aafi | 772 |  | 772 | 2.0% |
| Zarxio | filgrastim-sndz | 224 |  | 224 | 0.6% |
| Avasola | infliximab-axxq | 558 |  | 558 | 1.5% |
| Inflectra* | infliximab-dyyb | 3,648 | 270 | 3,918 | 10.3% |
| Ixifi* | infliximab-qbtx |  |  | - | 0.0% |
| Renflexis | infliximab-abda | 743 |  | 743 | 1.9% |
| Riabni | Rituximab-arrx | 567 |  | 567 | 1.5% |
| Ruxience | rituximab-pvvr | 614 |  | 614 | 1.6% |
| Truxima | rituximab-abbs | 1,179 |  | 1,179 | 3.1% |
| Subtotal |  | 29,270 | 1,034 | 30,304 | 79.4% |
| Biosimilar Drugs in Development |  |  |  |  |  |
| MYL-1701P | -- |  | 355 | 355 | 0.9% |
| TX05 | -- |  | 1,139 | 1,139 | 3.0% |
| BAT-1706 | -- | 760 |  | 760 | 2.0% |
| SB-8 | -- | 882 |  | 882 | 2.3% |
| AVT02 | -- | 1,031 | 548 | 1,579 | 4.1% |
|  |  |  |  | N | % |
| CHS-1420 | -- | 545 |  | 545 | 1.4% |
| FYB-201** | -- |  |  | - | 0.0% |
| SB11 | -- | 705 |  | 705 | 1.8% |
| MSB11455 | -- | 630 |  | 630 | 1.7% |
| TPI-120 | -- | 222 |  | 222 | 0.6% |
| *Subtotal* |  | 4,775 | 2,042 | 6,817 | 17.8% |
| **Failed Biosimilar Drugs** |  |  |  |  |  |
| Rixathon*** | rituximab | 1,048 |  | 1,048 | 2.7% |
| **TOTAL** |  | **35,093** | **3,076** | **38,169** | **100.0%** |

\* Inflectra was manufactured by Celltrion and marketed by Pfizer. Ixifi was manufactured/marketed by Pfizer. All infliximab biosimilar trials sponsored by Celltrion or Pfizer are listed under Inflectra.
\*\*Enrollment numbers were not listed on clinicaltrials.gov.
\*\*\*Rixathon received a Complete Response Letter from the FDA in 2018.

## Notes

### Competing Interest Statement

PBB reports grants from Kaiser Permanente, Arnold Ventures, advisory fees from EQRx, consulting fees from GRAIL and Foundation Medicine, speaking fees from Mercer, United Rheumatology, Morgan Stanley, Oppenheimer & Co, Cello Health, Oncology Analytics, Anthem, Magellan Health, Kaiser Permanente Institute for Health Policy, America's Health Insurance Plans, Geisinger, National Pharmaceutical Council, and stock from Grail, Oncology Analytics, EQRx. MRT reports LLC earnings distribution from Co-Bio Consulting, LLC, and speaking fees from Merck & Co and Shire.

### Author Declarations

As this study uses only publicly available non-individual-level data, it was exempt from IRB review.

